# Organized community sport participation in children and youth with physical disability: a scoping review protocol

**DOI:** 10.1101/2025.09.07.25335284

**Authors:** Karen Davies, Mitchell Barran, Colleen Pawliuk, Tiago Choi, Bruna Schilbach Pizzutto, Patricia Moreno Grangeiro, Courtney L. Pollock

## Abstract

Children and youth living with physical disability have less opportunities to participate in meaningful organized physical activities in their communities than their typically developing peers. Equitable access to community sport programs that aim to support the health, social participation, and well-being of these children and youth is required. Frameworks for program implementation and evaluation offer practical tools for supporting quality participation. Limited research exists demonstrating the practical application of these conceptual model frameworks. The objective of this scoping review is to understand the extent and type of evidence in relation to organized community sport participation in children and youth with physical disabilities. This review will consider studies that explore organized sport participation for children and youth up to 25 years with physical disability in all geographical areas. Children and youth with only a cognitive disability, developmental disability, or the absence of a physical disability will be excluded. The databases to be searched include MEDLINE (Ovid), Embase (Ovid), SportDiscus (EBSCO), CINAHL (EBSCO), PEDro, Global Index Medicus and Google Scholar. Sources of unpublished studies and grey literature will also be searched. No language or publication date restrictions will be placed on the search. Two independent reviewers will perform title and abstract screening, and retrieval and assessment of full-text citations. Any disagreements that arise between the reviewers at each stage of the selection process will be resolved through discussion or with a third reviewer. A narrative synthesis of the results will be presented in conjunction with a tabular and/or graphic format.

## Introduction

Every child has the right to participate in sports, yet it can be challenging for children and youth with physical disabilities to find meaningful sport participation opportunities in their community. The European Sports Charter defines sport as “ all forms of physical activity which, through casual or organized participation, are aimed at maintaining or improving physical fitness and mental well-being, forming social relationships or obtaining results in competition at all levels.” [1] Participation in sport provides health benefits and improves health outcomes for children and youth with physical disabilities,[2] promoting achievement of fine and gross motor skills, sport-specific physical ability, increased fitness, mobility, muscular strength, and opportunities for social network development.[3-5] Positive social experiences and meaningful connections enhance and promote participation in physical activity for youth with physical disabilities.[6] Despite these positive benefits and the well documented facilitators that enable sport participation for these children and youth, families and caregivers report a number of persistent environmental barriers. These include, but are not limited to, financial burden, transportation challenges, lack of information about existing programs, and physical accessibility.[6,7]

The World Health Organization describes disability as an inherent part of the human experience; the outcome of the interaction between an experienced health condition and/or impairment and circumstances associated with the environment and personal factors.[8] The minimum estimated global prevalence of children and adolescents younger than 20 years living with a disability is between 266 to 291.3 million (10.1-11.3%).[9] For youth ages 15-24 years, the prevalence is less clear. Children and youth with disabilities participate less in physical activity and recreation than their typically developing peers and are less likely to meet international guidelines for physical activity and sedentary behaviour.[5, 10-14] Further, children and adolescents with disability participate less, and with less diversity, frequency and intensity, in team or non-team sports when compared to their peers with typical development.[15,16] They are also more likely to play informal or casual games at home than more formal or organized sports in their community, even if sports are their preferred activities.[15,16] Limited evaluation of current community sport programs indicates the need for program implementation and evaluation frameworks to support quality participation for these children and youth.

The Canadian Disability Participation Project (CDPP) is a national organization that aims to improve quality participation in community-based sport and optimize outcomes of participation among children, youth and adults experiencing disability in collaboration with community sport partners.[17] They developed the Quality Parasport Participation Framework (QPPF) to identify quality experiences in sport using six building blocks: autonomy, belongingness, challenge, engagement, mastery, and meaning.[18,19] These building blocks are aligned with the barriers and facilitators of community-based sport participation for children and youth with physical disabilities and can be evaluated at the physical, program, and social environment levels. The RE-AIM Framework - <u>R</u>each for intended population; <u>E</u>ffectiveness of implementation; <u>A</u>doption by community; Implementation consistency, costs, adaptations; and <u>M</u>aintenance of intervention effects – has also been utilized to provide a comprehensive evaluation of public health programs.[20] Informed by this framework, Lawrason et al[21] have created a toolkit for applying the dimensions of RE-AIM in local community-based physical activity contexts for evidence-informed program evaluation.

A recent systematic review with meta-analysis identifying health outcomes for children and youth with physical disability participating in sports and physical recreation has identified several gaps in the literature.[2] They found that existing quantitative studies focused more on therapeutic and impairment models. Investigation of outcomes focused on participation occurring in community settings and more natural environments, to align with the goal of increasing health and inclusion for this population, is warranted.[2] Further, most reviews studied all physical activities together, leading to a lack of understanding regarding the effectiveness of activity in the individual therapy, exercise training, sport or recreation settings.[2] Primarily most population data describing physical activity participation in children and youth with disability also derives from high-income countries.[22] Evidence on the influence of physical activity among these children and youth remains limited. [2,22]

To our knowledge, no reviews have included a grey literature search on this topic to capture programs in middle- or lower-income countries and identify programs that already exist in communities around the world. A preliminary search of PROSPERO, MEDLINE, Open Science Framework and Google Scholar was conducted and no current or in-progress scoping reviews or systematic reviews on the topic were identified. Therefore, to further address the current gaps in the literature, the purpose of this scoping review is to systematically map and synthesize the level of evidence related to program evaluation of community sport participation globally in children and youth living with physical disability. A secondary aim is to further evaluate the perceived barriers and facilitators for community sport program implementation using framework evaluation.

## Review questions

1. What is the level of evidence related to program evaluation of community sport participation in children and youth up to 25 years of age with physical disabilities globally?
2. What are the perceived barriers and facilitators for community sport program implementation?

### Inclusion and exclusion criteria

#### Participants

This scoping review will consider studies that include children and youth up to 25 years of age with physical disabilities. Children and youth with only a cognitive disability, developmental disability, or the absence of a physical disability will be excluded. For this review, physical disability is defined broadly as the outcome of the interaction between a child or youth’s long-term physical health condition and their personal and environmental factors.[8]

#### Concept

This review will consider studies that explore organized sport participation for children and youth with physical disabilities. We will include studies with organized sport participation in a school setting but will exclude any school-based therapy interventions. Studies that describe adapted equipment, injury management, or para-athletics classification without addressing participation will be excluded. Unorganized sport, physical activity, and recreation studies will also be excluded.

#### Context

This review will consider studies from all geographic areas and will have a global scope.

#### Types of sources

This scoping review will consider quantitative, qualitative, and mixed methods study designs for inclusion. In addition, systematic reviews and un-published text and opinion papers will be considered for inclusion in the proposed scoping review. Protocols and/or clinical trial registries that do not include results will be excluded.

## Methods

The proposed scoping review will be conducted in accordance with the Joanna Briggs Institutes (JBI) methodology for scoping reviews [23] and will be reported in line with the Preferred Reporting Items for Systematic Reviews and Meta-Analyses extension for Scoping Reviews (PRISMA-ScR).[24] This protocol was developed in accordance with PRISMA for Protocols [25] (Supplement 1).

### Search strategy

The search strategy will aim to locate both published and unpublished primary studies. An initial limited search of MEDLINE (Ovid) and SportDiscus (EBSCO) was undertaken to identify articles on the topic. The text words contained in the titles and abstracts of relevant articles, and the index terms used to describe the articles, were used to develop a full search strategy for MEDLINE (Ovid; see Supplement 2). The search strategy, including all identified keywords and index terms, will be adapted for each included information source. The key words and index terms included were based on the experience of the authors and preliminary testing of the index terms. The reference lists of articles included in the review will be screened for additional papers using Web of Science Core Collection.

Articles from any language and those published from database inception to present will be included. We will search MEDLINE (Ovid), Embase (Ovid), SportDiscus (EBSCO), CINAHL (EBSCO), PEDro, Global Index Medicus and Google Scholar. Sources of unpublished studies and grey literature to be searched include: ProQuest Dissertations & Theses Citation Index (Web of Science), Open Access Theses and Dissertations (OATD), Papers First (WorldCat FirstSearch), Proceedings (WorldCat FirstSearch), PolicyCommons, Google, Biblioteca Virtual de Saúde (BVS) and Biblioteca Digital de Teses e Dissertações (BDTD).

### Study/Source of evidence selection

Following the search, all identified records will be collated and uploaded into Covidence (Veritas Health Innovation, Melbourne, Australia) and duplicates removed. Following a pilot test, titles and abstracts will then be screened by two independent reviewers for assessment against the inclusion criteria for the review. Potentially relevant papers will be retrieved in full. The full text of selected citations will be assessed in detail against the inclusion criteria by two independent reviewers. Reasons for exclusion of full-text papers that do not meet the inclusion criteria will be recorded and reported in the scoping review. Any disagreements that arise between the reviewers at each stage of the selection process will be resolved through discussion or with a third reviewer. The results of the search will be reported in full in the final scoping review and presented in a PRISMA flow diagram.[26]

### Data extraction

Data will be extracted from papers included in the scoping review by two independent reviewers using a data extraction tool developed by the reviewers. The data extracted will include specific details about the study design and methods, participants, sport and context characteristics, outcomes defined by the RE-AIM and QPPF frameworks, barriers and facilitators of implementation, integration models, and other key elements relevant to the review questions. A draft extraction tool is provided (see Supplement 3). The draft data extraction tool will be modified and revised as necessary during the process of extracting data from each included paper. Modifications will be detailed in the full scoping review. Any disagreements that arise between the reviewers will be resolved through discussion or with a third reviewer. Authors of papers will be contacted to request missing or additional data, where required.

### Data analysis and presentation

Two frameworks will guide the critique of studies found in the scoping review; 1) RE-AIM Framework;[20] Reach of the program; Effectiveness of implementation; Adoption by community members; Implementation of support strategies; and Maintenance of success and and 2) the Quality Parasport Participation Framework (QPPF).[17] We will share a narrative synthesis of the results and tables and/or graphs as appropriate in accordance with the review objectives and questions.

## Data Availability

All relevant data from this study will be made available upon study completion.

## Supplement 1: PRISMA for Protocols Checklist

## Supplement 2: Search Strategy

Ovid MEDLINE(R) ALL <1946 to june 06, 2025>

| # | Query | Results from<br>9 Jun 2025 |
| --- | --- | --- |
| 1 | Children with Disabilities/ | 7,447 |
| 2 | (disabil* or impairment?).tw,kf. | 733,740 |
| 3 | Cerebral Palsy/ | 25,262 |
| 4 | (cerebral palsy or spastic diplegia).tw,kf. | 30,759 |
| 5 | Meningomyelocele/ | 4,726 |
| 6 | (meningomyelocele or spina bifida).tw,kf. | 10,023 |
| 7 | (neurological adj2 (diseas* or disab* or disorder?)).tw,kf. | 80,202 |
| 8 | Brain Injuries, Traumatic/ | 16,359 |
| 9 | acquired brain injur*.tw,kf. | 3,595 |
| 10 | exp Limb Deformities, Congenital/ | 27,236 |
| 11 | (limb deformit* or limb deficienc*).tw,kf. | 2,012 |
| 12 | exp Joint Diseases/ | 457,092 |
| 13 | Legg-Calve-Perthes Disease/ | 2,053 |
| 14 | "Developmental Dysplasia of the Hip"/ | 638 |
| 15 | (joint disease? or joint deficienc* or Legg-Calve-Perthes Disease or "Developmental Dysplasia of the Hip" or congenital hip dysplasia or developmental hip dysplasia).tw,kf. | 20,445 |
| 16 | Spinal Cord Injuries/ | 47,397 |
| 17 | (spinal cord Injur* or post-traumatic myelopathy).tw,kf. | 53,207 |
| 18 | or/1-17 | 1,383,197 |
| 19 | (sports/ or baseball/ or basketball/ or bicycling/ or boxing/ or cricket sport/ or football/ or gaelic football/ or golf/ or | 47,771 |
|  | gymnastics/ or hockey/ or martial arts/ or tai ji/ or mountaineering/ or racquet sports/ or tennis/ or return to sport/ or rugby/ or running/ or jogging/ or marathon running/ or skating/ or snow sports/ or skiing/ or soccer/ or sports for persons with disabilities/ or team sports/ or lacrosse/ or "track and field"/ or volleyball/ or walking/ or nordic walking/ or water sports/ or swimming/ or diving/ or weight lifting/ or wrestling/ or youth sports/) and (community or adapt* or organized or program* or modif* or para or intervention?).tw,kf. |  |
| 20 | ((sport? or sporting or athletics or baseball or basketball or bicycling or cycling or boxing or cricket or football or golf or gymnastics or hockey or martial arts or tai ji or tai chi or hiking or tennis or rubgy or running or jogging or skating or skiing or snowboarding or lacrosse or "track and field" or volleyball or walking or swimming or diving or rowing or weight lifting or strength training or wrestling or mobility or stretching) adj5 (community or adapt* or organized or program* or modif* or para or intervention?)).tw,kf. | 29,955 |
| 21 | 19 or 20 | 66,589 |
| 22 | participat*.tw,kf. | 770,851 |
| 23 | exp child/ or exp infant/ or adolescent/ or exp pediatrics/ or child, abandoned/ or exp child, exceptional/ or child, orphaned/ or child, unwanted/ or minors/ or young adult/ or (pediatric* or paediatric* or (child* not childbearing) or newborn* or congenital* or infan* or baby or babies or neonat* or pre-term or preterm* or premature birth* or NICU | 5,994,145 |
|  | or preschool* or pre-school* or kindergarten* or kindergarden* or elementary school* or nursery school* or (day care* not adult*) or schoolchild* or toddler* or boy or boys or girl* or middle school* or pubescen* or juvenile* or teen* or youth* or young adult? or young people or high school* or adolesc* or pre-pubesc* or prepubesc*).mp. or (child* or adolesc* or pediat* or paediat*).jn. |  |
| 24 | 18 and 21 and 22 and 23 | 916 |

## Supplement 3: Data Extraction Instrument

The chart headings to extract data are as follows:

Authors, year of publication, and country of origin

Study title, aims, design, and duration

Research question or objective, measured outcome, and tools of measurement

Implementation framework or integration model

Study population and total number of participants

Organized sport type, frequency, and duration

Identified facilitators and barriers of implementation

Summary of findings

